# Relationship between eye movements and freezing of gait during turning in individuals with Parkinson’s disease

**DOI:** 10.1101/2021.09.06.21263180

**Authors:** Ying Wang, Marc M. van Wanrooij, Rowena Emaus, Jorik Nonneke, Michael X Cohen, Helena Cockx, Sabine Janssen, Rick C. Helmich, Richard van Wezel

**Affiliations:** Department of Biophysics, Donders Institute for Brain, Cognition and Behaviour, Radboud University, the Netherlands; Department of Electrical Engineering, Eindhoven University of Technology, Eindhoven, the Netherlands; Academic Center for Epileptology Kempenhaeghe, Heeze, the Netherlands; Radboud University Medical Centre; Donders Institute for Brain, Cognition and Behaviour; Department of Rehabilitation; Centre of Expertise for Parkinson & Movement Disorders, Nijmegen, The Netherlands; Department of Rehabilitation, Sint Maartenskliniek, Nijmegen, the Netherlands; Donders Centre for Medical Neuroscience, Radboud University Medical Center, Nijmegen; Department of Neurology, Donders Institute for Brain, Cognition and Behaviour, Radboud University Medical Centre, Nijmegen, the Netherlands; Biomedical Signals and Systems Group, Faculty of Electrical Engineering, Mathematics and Computer Science (EEMCS), University of Twente, Enschede, the Netherlands

**Keywords:** Turning, eye movement, electrooculography, gaze, FOG

## Abstract

**Background:** Individuals with Parkinson’s disease can experience freezing of gait: “a sudden, brief episode of an inability to move their feet despite the intention to walk”. Since turning is the most sensitive condition to provoke freezing-of-gait episodes, and the eyes typically lead turning, we hypothesize that disturbances in saccadic eye movements are related to freezing-of-gait episodes.

**Objectives:** This study explores the relationship between freezing-of-gait episodes and saccadic eye movements for gaze shift and gaze stabilization during turning.

**Methods:** We analyzed 277 freezing-of-gait episodes provoked among 17 individuals with Parkinson’s disease during two conditions: self-selected speed and rapid speed 180-degree turns in alternating directions. Eye movements acquired from electrooculography signals were characterized by the average position of gaze, the amplitude of gaze shifts, and the speed of gaze stabilization. We analyzed these variables before and during freezing-of-gait episodes occurring at the different phase angles of a turn.

**Results:** Significant off-track changes of the gaze position were observed almost one 180-degree-turn time before freezing-of-gait episodes. In addition, the speed of gaze stabilization significantly decreased during freezing-of-gait episodes.

**Conclusions:** We argue that off-track changes of the gaze position could be a predictor of freezing-of-gait episodes due to continued failure in movement-error correction or an insufficient preparation for eye-to-foot coordination during turning. The decline in the speed of gaze stabilization is large during freezing-of-gait episodes given the slowness or stop of body turning. We argue that this could be evidence for a healthy compensatory system in individuals with freezing-of-gait.

## INTRODUCTION

Freezing of gait is a symptom commonly observed in the moderate to advanced stages of Parkinson’s disease and defined as “brief, episodic absence or marked reduction of forward progression of the feet despite the intention to walk.” ^1^. Turning is typically considered to be difficult for individuals with Parkinson’s disease and to be the most sensitive trigger of freezing-of-gait episodes ^2,3^. The unpredictable occurrences of freezing-of-gait episodes during turning lead to a high risk of falls and may cause fall-related fractures ^4^.

Eye movements play an important role in turning which requires complex eye-foot coordination. The eyes lead the head and body rotation through rhythmic and involuntary oscillations during turning. In further detail, saccadic eye movements shift the gaze to the direction of turning (gaze shift) and stabilize the gaze direction (gaze-direction stabilization) through vestibular-ocular reflex to compensate for rapid head rotation ^5–7^. Eye movements become crucial during turning in individuals with mild Parkinson’s disease ^8^, and individuals with Parkinson’s disease use larger eye movements in the eye-foot coordination compared to healthy controls ^8^. The use of larger eye movements is most likely a visual compensatory strategy for the movement slowness of the head and body in Parkinson’s disease ^8,9^. With disease progression, the ocular-motor system becomes gradually impaired, and this may drive the eyes way off target ^10^: errors in shifting gaze to correct turning directions and/or errors in stabilizing the gaze direction during head rotation. Failure of this ocular compensatory mechanism may result in defective turning performance ^11^, such as freezing-of-gait episodes.

Individuals with a history of frequent freezing-of-gait episodes have an impaired ocular-motor system with great variability of saccade velocity and gain ^12^. However, direct evidence linking saccadic eye movements to freezing-of-gait episodes is missing. Here, we explored gaze shifts and gaze-direction stabilizations before and during freezing-of-gait episodes evoked by turning in individuals with Parkinson’s disease. Given that gaze shifts guide the head and body rotation during turning, we hypothesized that the individuals keep correct gaze shifts until the onsets of freezing-of-gait episodes. Furthermore, we assumed that individuals with Parkinson’s disease have a healthy vestibulo-ocular reflex system. The disappearance of gaze-direction stabilization during freezing-of-gait episodes is hypothesized because the vestibular-ocular system is relatively insensitive to slow rotation of the head ^7^, and individuals with Parkinson’s disease move their body slowly or even stop moving during freezing-of-gait episodes.

To visualize the expected gaze shift and stabilization of gaze direction before and during freezing-of-gait episodes, we present the illustration of our hypotheses in *figure 1* where x-axis indicates time relative to freezing onset. The body-turning position of participants is ideally simulated as sinus waves (*figure 1A*). Gaze shifts and gaze-direction stabilizations which are elicited by the body turning are called quick-phase and slow-phase eye movements, respectively. The quick-phase eye movements are described by two variables: center-of-beat which indicates the average gaze position during a gaze shift and quick-phase amplitude indicates the amplitude of a gaze shift. The slow-phase eye movements are described by slow-phase velocity which indicates the speed of motion of the visual image on the retina surface to counteract the head rotation. The center-of-beat and the quick-phase amplitude (*figure 1B*), and the slow-phase velocity (*figure 1C*) were calculated from the simulated body-turning position.

**Figure 1.**
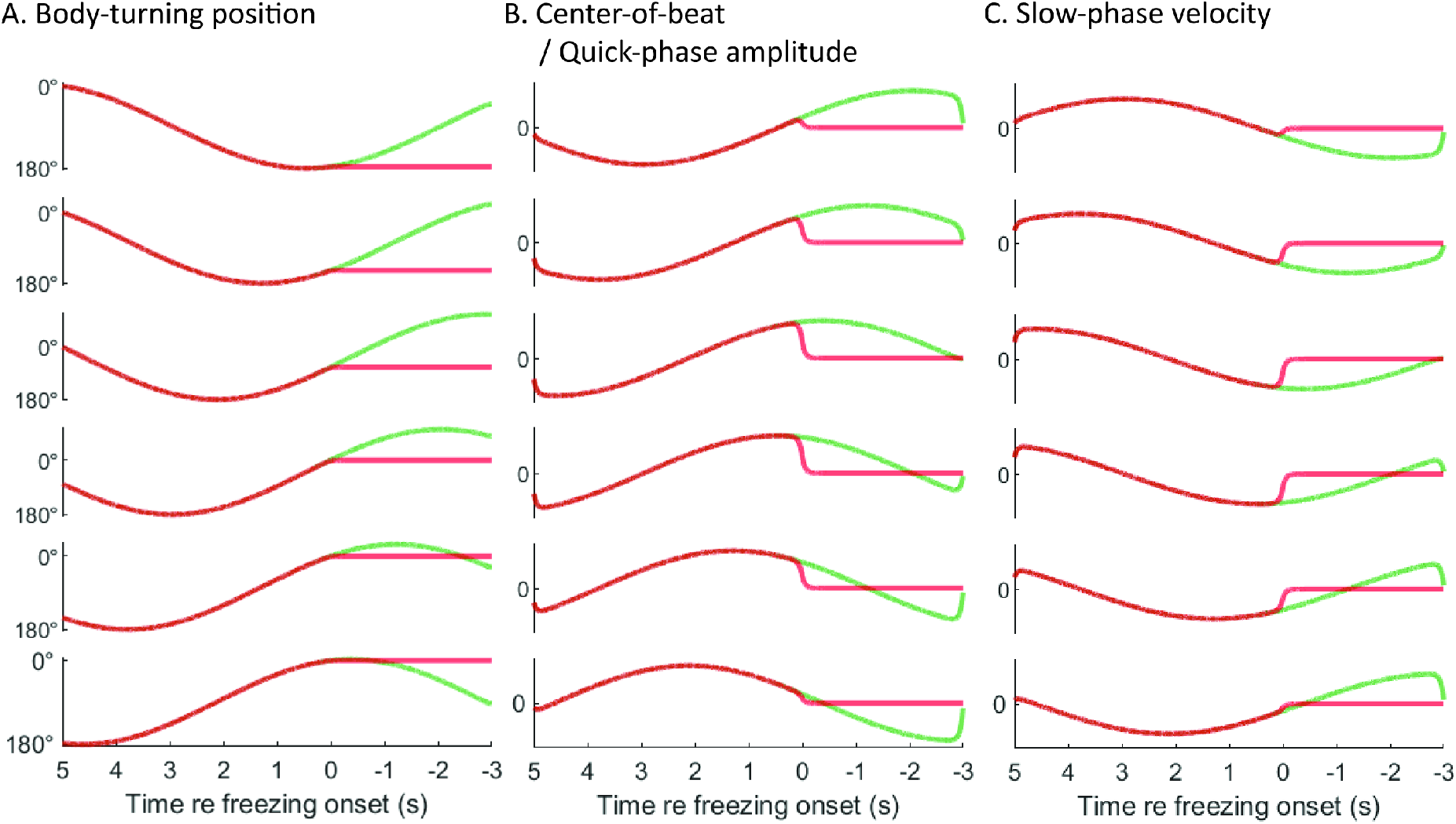
Illustration of null hypotheses: individuals keep correct gaze shift until freezing-of-gait onsets, and gaze-direction stabilizations disappear during freezing-of-gait episodes. Figure 1A shows that the participants execute 180-degree turns in alternating directions, and the six rows in the figure indicate that freezing episodes occur at six turn-phase-angle sections: 0-30 degree, 30-60 degree, 60-90 degree, 90-120 degree, 120-150 degree, and 150-180 degree relative to the start of turning. The red and green curves indicate turning affected and not affected by freezing episodes, respectively. 0 s indicates the onsets of freezing episodes. The illustration shows that the saccadic eye movements—center-of-beat and quick-phase amplitude (B), and slow-phase velocity (C)—are close to zero when participants do not change their body position during turning (A) because of freezing episodes. The body position during turning was ideally simulated as sinus curves, and the duration of each 180-degree turn was modeled as 5 seconds. The center-of-beat and quick-phase amplitude, and slow-phase velocity were defined as the first derivative of the body position but with different directions. The center-of-beat and the quick-phase amplitude have the same direction as the body turning because these two variables describe the gaze shift for guidance during turning. The slow-phase velocity describes the gaze-direction stabilization during turning and therefore has the opposite direction as the turning body.

## METHOD

### Participants

Seventeen participants with idiopathic Parkinson’s disease were recruited. We included participants that experienced regular freezing-of-gait episodes (“very often, more than one time a day” in the freezing frequency section of the New Freezing of Gait Questionnaire ^13^) in the past month. We excluded participants with comorbidities that cause severe gait impairment, severe cognitive impairments (the score of a Mini-Mental State Examination ^14^ <24), and inability to walk 150 meters unaided.

### Ethical approval

This study was ethically approved by the Dutch committee on research involving human participants [Arnhem-Nijmegen region (NL60942.091.17)]. The experiment conformed to the Declaration of Helsinki, and all participants provided written informed consent.

### Setup

Four electrooculography electrodes (active electrodes from actiCAP, Brain Products GmbH) were aligned with the center of pupils in the vertical and horizontal planes when the participants looked straight ahead. To increase the quality of data, viscous gels (Supervisc) were applied to bridge the electrodes and the skin. In this study, the electrooculography signals (an example in *figure 2A*) had a sampling rate of 500 Hz.

**Figure 2.**
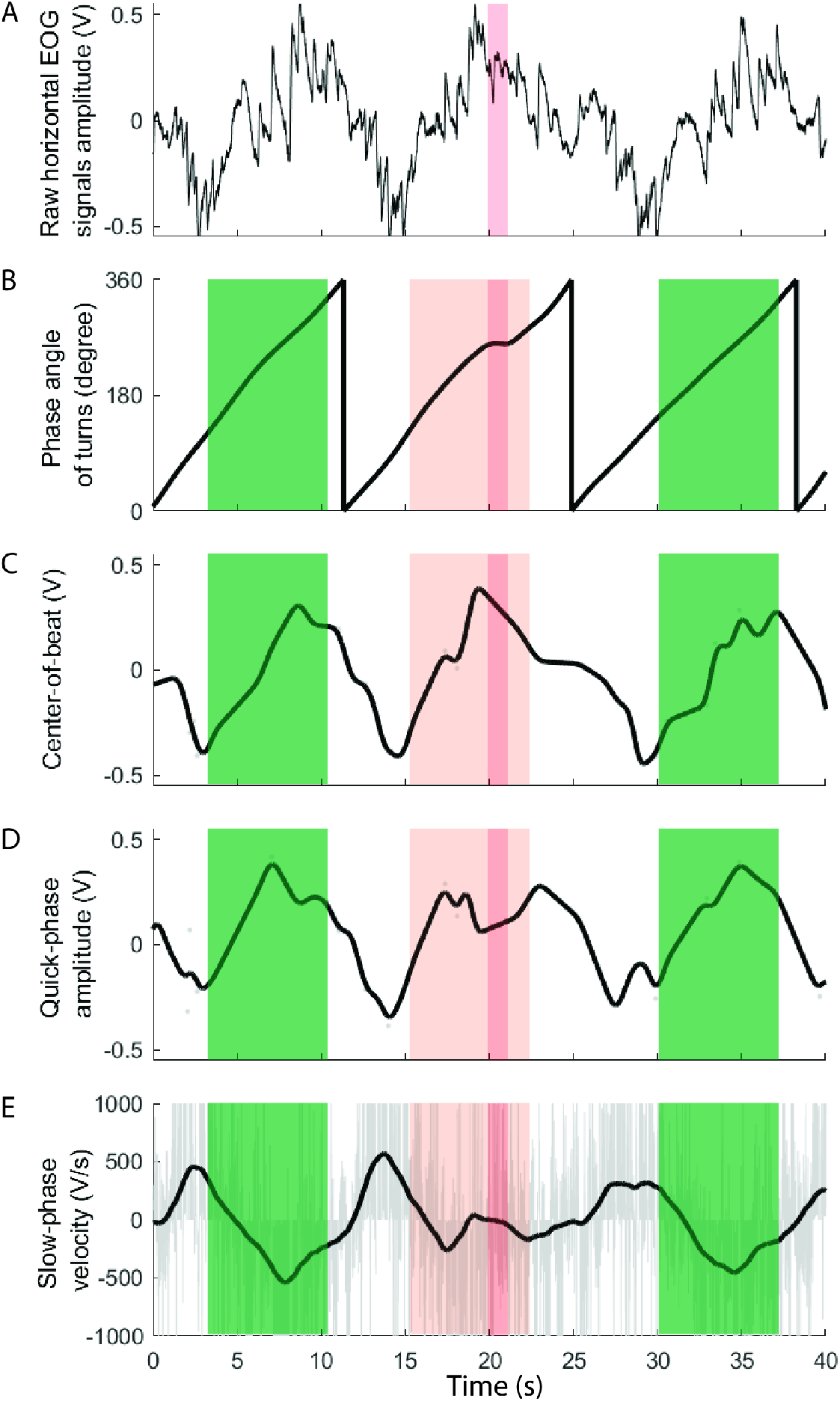
Illustration of the electrooculography signal analysis: An example (A) of raw electrooculography signals, the phase angle of turning (B), and the variables of saccadic eye movements—center-of-beat (C), and quick-phase amplitude (D), and slow-phase velocity (E). The 0-180 degree in the phase angle of turning (B) indicates the clockwise 180-degree turns, and the 180-360 degree indicated the counterclockwise turns. The red patches in *figure 2A* indicate the freezing episodes. The red patches in *figure B-E* indicate the eye movements before and during freezing of gait, and the green patches indicate the eye movements during turns which were not affected by freezing episodes, respectively. The red and green segments have the same phase angles at their start for the comparison of eye movements in the two categories.

### Paradigm

Participants were assessed in the dopaminergic OFF-medication state, after 12 hours of dopaminergic medication withdrawal. The data were collected when the participants executed two turn conditions ^15^: alternating half turns—clockwise and counterclockwise 180-degree turns—at a self-selected normal speed and a rapid speed. Each condition lasted 2 minutes, and participants repeated each condition maximally five times.

### Annotation of freezing of gait

All experiments were video-taped with two video cameras: a GoPro Hero5 and a Sony-HDR video-camera. The cameras were placed in the front and on the side of the participants for precise freezing-of-gait annotations, respectively. The freezing-of-gat annotations were mainly based on the videos recorded by the camera in the front (GoPro), and the videos from the camera on the side (Sony-HDR) assisted the raters on uncertain freezing-of-gait episodes. Freezing-of-gait episodes and unexpected movements during the sessions, such as sudden stops caused by other reasons than freezing, were annotated by two independent experienced raters using an open-source annotation software (ANVIL ^16^). When the two raters disagreed on some annotations, a third experienced rater made the final decision.

### Eye movement preprocessing

We calculated three variables from the horizontal electrooculography signals: the center-of-beat, the quick-phase amplitude, and the slow-phase velocity. The amplitude of the horizontal electrooculography signals was calculated by subtracting the signals of the right electrooculography electrode from the signals of the left electrode. The center-of-beat was calculated as the average amplitude of the horizontal electrooculography signals during quick-phase eye movements. The quick-phase amplitude was calculated as the amplitude difference between the onset and offset of quick-phase eye movements. The slow-phase velocity was calculated as the first derivate of the amplitude of the horizontal electrooculography signals during slow-phase eye movements.

We identified the time stamps of the quick-phase and slow-phase eye movements through a customizable algorithm. A detailed workflow of the algorithm was presented in *appendix A* based on the Cluster Fix ^17^ saccade detection algorithm. We firstly applied the Cluster Fix algorithm to partition electrooculography signals into two clusters: quick-phase and slow-phase eye movements. Given that some of the detected quick-phase eye movements look like slow-phase eye movements which may affect the analysis of variables for quick-phase eye movements, we further refine the cluster of quick-phase eye movements through K-means clustering with three features of main sequence for human saccades ^18^: amplitude, duration, and velocity. We visually checked the conservative quick-phase eye movements and ensured that no slow-phase eye movements were included in the quick-phase cluster, and some quick-phase eye movements similar as the slow-phase eye movements were probably excluded because of the strict criterion.

### Eye movement analysis in turns

To explore freezing-of-gait episodes occurring at different turn phases, we divided each 180-degree turn into six sections—0-30 degrees, 30-60 degrees, 60-90 degrees, 90-120 degrees, 120-150 degrees, and 150-180 degrees. The freezing-of-gait episodes were assigned to the individual sections based on the instantaneous phase angles of the freezing-of-gait onsets. The instantaneous phase angles of the turns were estimated from the analytic signals of the slow-phase velocity using the Hilbert transformation^19^ because the slow-phase velocity is the indicator for gaze stabilization during turning. In addition, we assumed no relative difference among the eye movements in clockwise and counterclockwise turns, therefore the eye movements started during the clockwise (*figure 1A, top*: start turning from 0 degree to 180 degree) and counterclockwise (*figure 1A, bottom*: start turning from 180 degree to 0 degree) turns were merged through flipping over the eye movements during counterclockwise turns. Furthermore, the duration of each 180-degree turn was unified as 5 seconds given that the durations of individual turns were different. In other words, the phase angle of turning at the 5^th^ second before freezing-of-gait onsets was as same as the angle of the freezing-of-gait onsets.

We divided the eye movements into two categories—no-freezing turning and freezing turning—to investigate eye movements during turns that were assumed to be not affected and affected by freezing-of-gait episodes, respectively. An illustration is shown as green and red patches in *figure 2 B-E*. We assumed that eye movements earlier than one-turn time before freezing-of-gait onsets (earlier than the 5^th^ second before freezing-of-gait onsets) are not related to freezing-of-gait episodes. Given that most freezing-of-gait episodes are generally shorter than 3 seconds, eye movements within the period from the 5^th^ second before to the 3^rd^ second after freezing-of-gait onsets were segmented and added in the category of freezing turning. Moreover, each segment only included one freezing-of-gait episode to avoid the influences of other freezing-of-gait episodes. The eye movements in the no-freezing-turning category were segmented corresponding to each segment in the freezing-turning category with the same segment length and segment-onset phase angle. Each segment in the freezing-turning category may have multiple corresponding segments in the no-freezing-turning category, which were collected from the same experiment condition.

The means of the eye-movement segments in the no-freezing-turning and freezing-turning categories were compared with Student t-tests. To explore the eye movements with the evolution of freezing-of-gait episodes, we visualized the t-test through plotting the mean and its 95% confidence intervals of the eye-movement variables at each sampling point. When there is no overlap between the plots of the center-of-beat in the two categories before a freezing-of-gait episode (from the 5^th^ second before freezing-of-gait onsets to the freezing-of-gait onsets), our first hypothesis—individuals keep correct gaze shifts until freezing-of-gait onsets—is rejected. When there is a considerable overlap between the plots of the slow-phase velocity in the two categories during freezing-of-gait episodes (from freezing-of-gait onsets to the 3^rd^ second after the freezing-of-gait onsets), our second hypothesis—gaze-direction stabilizations disappear during freezing-of-gait episodes in individuals with Parkinson’s disease—is rejected. In addition, given that the quick- and slow-phase eye movements consecutively present during turning to shift the gaze and compensate the gaze direction, the center-of-beat, quick-phase amplitude, and slow-phase velocity were discontinuous; hence, these variables were linearly interpolated for the visualization.

## RESULTS

### Participants

Fifteen of the 17 participants’ data were successfully collected. The median age of the 15 participants was 73 years in the 51-89 age range. The median disease duration was 12 years (range: 3-20 years). The clinical characteristics of the participants were assessed by several clinical examinations and questionnaires and shown in *Table 1*. According to the answers in New Freezing of Gait Questionnaire, all participants except one experienced freezing-of-gait episodes during turning in daily life, and eight of them experienced “very often, more than once a day” freezing during turning.

**Table 1.** Clinical characteristics of participants.

| Clinical examinations and questionnaires | Scores: minimum to maximum (median) |
| --- | --- |
| Movement Disorders Society-Unified Parkinson’s disease Rating Scale Part III <sup>25,26</sup> | 24 to 49 (32) |
| Hoehn and Yahr stage <sup>27</sup> | 2 to 4 (2) |
| New Freezing of Gait Questionnaire <sup>13</sup> | 10 to 25 (20) |
| Mini-Mental State Examination <sup>14</sup> | 24 to 30 (29) |
| Frontal Assessment Battery <sup>28</sup> | 14 to 18 (17) |

### Freezing of gait

In total, 511 freezing-of-gait episodes were annotated, and the median duration of the episodes was three seconds (range: around one second to two minutes). The median number of episodes across patients was 28 in the range of 12 to 101. A substantial interrater agreement for the annotation of freezing-of-gait episodes was reached with around 92% in percent-agreement and 0.78 in Cohen’s kappa. According to the criteria in the eye-movement-analysis section, 277 segments were included in the category of freezing turning. The number of segments were 69, 55, 36, 36, 24, and 57 when freezing-of-gait episodes occurred within the six turn-phase-angle sections from 0 to 180 degrees (details are shown on the left side of *figure 3*). This finding supports the previous studies that freezing-of-gait episodes commonly occur when the individuals initiate turns and approach destinations ^20^.

**Figure 3.**
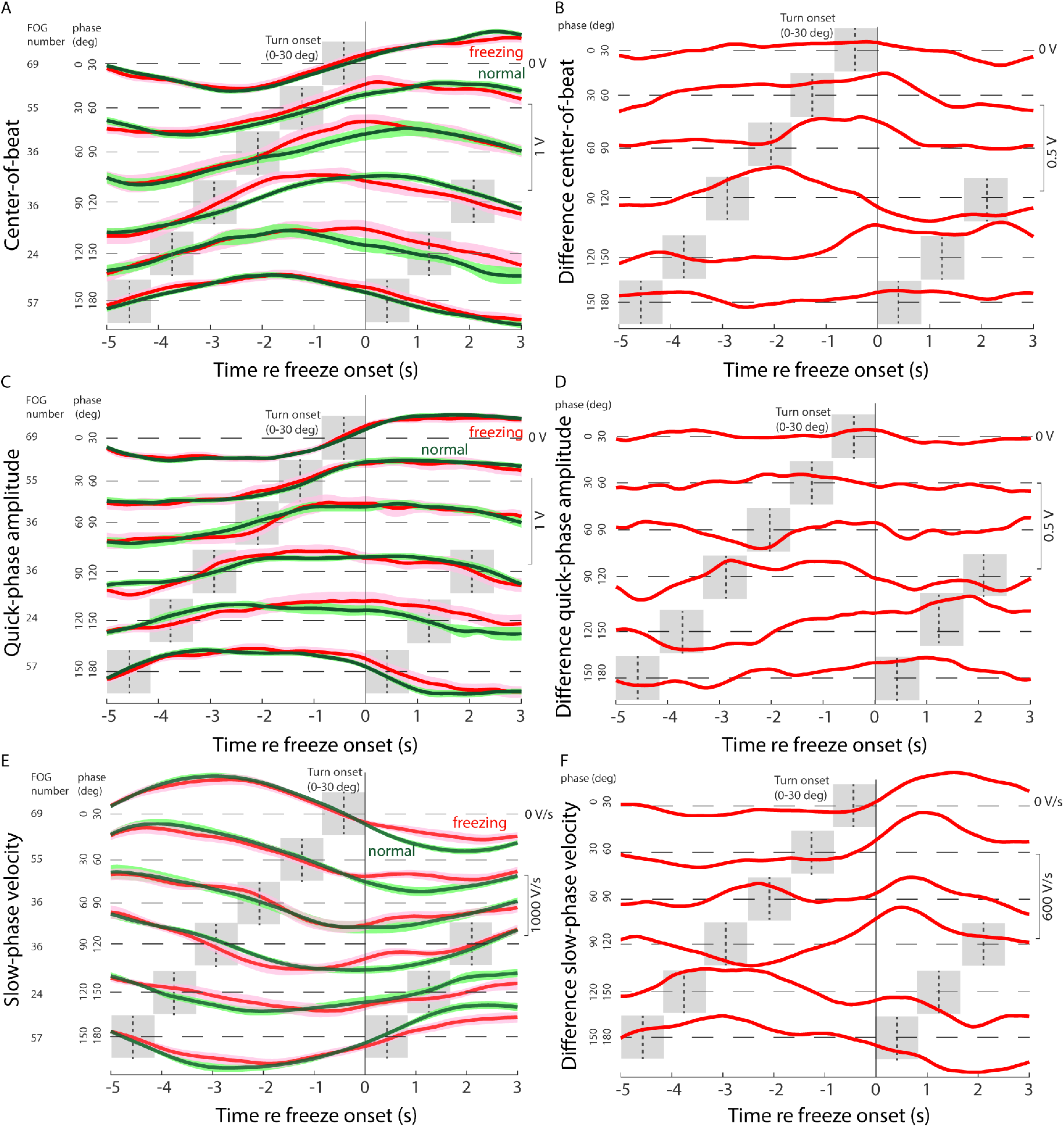
Comparison between the eye movements in two categories: freezing-turning (“freezing” in the figure; red curves) and no-freezing-turning (“normal” in figure; green curves). The eye-movement variables are center-of-beat (*figure A and B*), quick-phase amplitude (*figure C and D*), and slow-phase velocity (*figure E and F*). The 0 s indicates the onsets of freezing episodes. There are six rows indicating freezing episodes occurred within 0-30 degree, 30-60 degree, 60-90 degree, 90-120 degree, 120-150 degree, and 150-180 degree, respectively. The curves are the mean, and the patches are the 95% confidence intervals of the mean in *figure A, C*, and *E*. The curves in *figure B, D*, and *F* indicate the difference between the eye movements in the two categories. The gray patches indicate the start of a new turn.

### Eye movements

*Figure 3* presents the t-test visualization of the eye movement variables: the center-of-beat (*figure 3A*), the quick-phase amplitude (*figure 3C*), and the slow-phase velocity (*figure 3E*). To further explore trends in the eye-movement variables before and during freezing-of-gait episodes, we calculated the differences between the no-freezing-turning and freezing-turning categories through subtracting the mean of variables in the no-freezing-turning category from the mean in the freezing-turning category. The differences are shown in *figure 3B, D*, and *F*.

The center-of-beat significantly changed before the freezing-of-gait onsets (*figure 3A*). As shown in *figure 3B* (the zero-crossing points in the sections of 0-120 degree), the center-of-beat in the freezing-turning category differed no-freezing-turning category even 4 seconds (almost the duration of one turn) before freezing-of-gait onsets; in other words, participants started looking somewhere else than a correct turning orbit before freezing-of-gait episodes. Of note, the difference increased after the start of a new turn (indicated by gray patches in *figure 3*), and the difference became larger when the duration between the start of a new turn and freezing-of-gait onsets was longer. Furthermore, given that the changes of turning body are guided by the center-of-beat, significant changes in the slow-phase velocity were expected to be observed before freezing-of-gait onsets (*figure 3E*). Interestingly, the slow-phase velocity changed slightly earlier than the center-of-beat as shown in *figure 3B* and 3*F*. In addition, the corresponding changes of the quick-phase amplitude were also observed in *figure 3C* given that the quick-phase amplitude is a variable related to both center-of-beat and slow-phase velocity.

As shown in *figure 3E* and *3F*, the magnitude of slow-phase velocity significantly decreased and was close to zero during freezing-of-gait episodes. The magnitude of the center-of-beat (*figure 3A* and *3B*) and quick-phase amplitude (*figure 3C* and *3D*) decreased during freezing-of-gait episodes, but the decrease was not significant as the slow-phase velocity. The decreases of the center-of-beat and quick-phase amplitude were probably limited in time, and any clear effect was obfuscated given that participants tried to reach the end of a turn or restart turning during freezing-of-gait episodes.

## DISCUSSION

This study offers some essential insights into the temporal relationships between the saccadic eye movements and freezing-of-gait episodes provoked by turning conditions for individuals with Parkinson’s disease. Three eye-movement variables—center-of-beat, quick-phase amplitude, and slow-phase velocity—were analyzed to investigate the performance of shifting gaze to turning direction before freezing-of-gait-episode onsets and the performance of stabilizing gaze direction to compensate for head rotation during freezing-of-gait episodes. Significant differences in center-of-beat were found in the duration of almost one turn before freezing-of-gait episodes. This implies that our first hypothesis is rejected, and individuals look somewhere else than at the correct turning direction. During freezing-of-gait episodes, the magnitude of slow phase velocity significantly decreased, and our second hypothesis—gaze-direction stabilizations disappear during freezing-of-gait episodes in individuals with Parkinson’s disease—is not rejected. This could be evidence that individuals with Parkinson’s disease and freezing-of-gait episodes have a healthy compensatory vestibulo-ocular reflex.

The changes of center-of-beat in the orbit of a normal turn before freezing-of-gait episodes could be caused by several possible factors. (1) Abnormal gaze-direction stabilization, which may be caused by the dysfunction of striato-nigro-collicular pathway in Parkinson’s disease ^10^, possibly lead to abnormal gain of retinal slip which can drive the gaze get off the track of the correct turning direction ^10^. This may also explain why the slow-phase velocity changed earlier than the center-of-beat before freezing-of-gait episodes in *figure 3B* and *F*. (2) The decreased inhibition of reflexive saccades and attention deficits in individuals with Parkinson’s disease ^21^ could also result in the off-track gaze direction during turning. In this study, we did not formally test the presence of attention deficits although the presence of cognitive impairments was an exclusion criterion. We recommend further investigation on the possible factors of the center-of-beat changes in future research.

The beginning of a new turn can worsen the off-track problem and even result in freezing-of-gait episodes. As shown in *figure 3B*, after an unexpected off-track error occurs, the error is most likely kept and even enlarged by the start of a new turn until a freezing-of-gait episode is provoked. This finding also supports other earlier research which observes that initial saccades of a turn can predict turning performance in individuals with Parkinson’s disease ^11^.

The significant off-track changes of the center-of-beat before the freezing-of-gait onsets could be speculated as a predictor of freezing-of-gait episodes evoked by turning. One possible explanation for this might be that individuals may keep correcting the off-track gaze direction during turning, but the correction failed because of a deficit in motor planning in individuals with freezing-of-gait episodes ^12,22^. Accordingly, the continued failure in motor-error correction might cause freezing-of-gait episodes. Another possible explanation is that an unexpected large gaze shift, that is, individuals with Parkinson’s disease look at faraway objects, could result in insufficient anticipatory coordination between the body segmentations for turning which most likely provokes freezing-of-gait episodes ^8^. The unexpected large shift could be avoided by visual cues, which guides individuals to turn at a suitable amplitude and speed ^23^.

This study has several limitations. Given that this is an explorative study, we only included 17 participants and measured the physiological signals from 15 of the 17 participants at off-medication state to acquire sufficient freezing-of-gait episodes for the analysis. The small number of participants may limit the generalization of the interpretation about the relationship between the eye movements and freezing-of-gait episodes evoked by turning. Future work should include more participants and collect the signals at on-medication state to examine how dopaminergic treatment affects the eye movements during freezing-of-gait episodes. In addition, we assumed no considerable difference of the saccadic eye movements between clockwise and counterclockwise turns, but the differences may exist among individuals due to the asymmetric motor function in patients with Parkinson’s disease ^24^. In future studies, researchers could investigate whether the eye movements in clockwise and counterclockwise turns are different and what factors contribute to the differences using for example the asymmetry assessment scores in Movement Disorders Society-Unified Parkinson’s disease Rating Scale Part III. In this study, we did not collect gyroscope signals for the turn angle of the head and body, and only did horizontal saccadic analysis during turning using electrooculography signals, but the relationship between the eye-foot coordination and freezing-of-gait episodes would be valuable to be determined in future research. Researchers in following studies are recommended to use more advanced technical knowledge and devices, e.g., head-body motion sensing systems for the analysis of the eye-to-foot coordination, and video-oculography for the reliable identification and analysis of the other types of eye movements especially during natural tasks in daily life, such as blinks and smooth pursuit movements. Studies that investigate dynamic eye movements with visual cues as a sensory compensation strategy to overcome freezing-of-gait episodes would be helpful to determine how the visual cues work on the eye movements for alleviating freezing-of-gait episodes. Furthermore, multimodal physiological signals for the brain, eye, heart activity, and gaits can be simultaneously analyzed for a deep and comprehensive understanding about freezing-of-gait episodes in our future works.

## Conclusion

In this study, we investigated the relationship between the saccadic eye movements and freezing-of-gait episodes evoked by turning in individuals with Parkinson’s disease. Individuals were found to look at a different direction from the correct turning direction before freezing-of-gait episodes, which can be a predictor for freezing-of-gait episodes. Furthermore, we found that the magnitude of slow phase velocity significantly decreased which could be an evidence that individuals with Parkinson’s disease and freezing-of-gait episodes have a healthy compensatory vestibulo-ocular reflex.

## Data Availability

Anonymous data sets referred to this manuscript can be shared through the contact of corresponding author.

## Acknowledgments

We are grateful to all participants for their time and effort. This work was strongly supported by Dutch Parkinson’s disease Association (Parkinson Vereniging). We especially thanks Jan Gouman, Piet van de Schilde, and Erik Jan Marinissen for their precious suggestions. Besides, we are also grateful to the contribution during the experiments and data analysis from the assistants: Karlijn van Dijsseldonk, Floris Beuving, Rowena van der Velden, Günter Windau, and Jessica Askamp.

## Author Roles

*1. Research project: A. Conception, B. Organization, C. Execution; 2. Statistical Analysis: A. Design, B. Execution, C. Review and Critique; 3. Manuscript Preparation: A. Writing of the first draft, B. Review and Critique;*

Ying Wang contributed to 1A, 1B, 1C, 2A, 2B, 2C, 3A, and 3B.

Marc M. van Wanrooij contributed to 1A, 2A, 2B, 2C, and 3B.

Rowena Emaus contributed to 1C, 2B, and 3B.

Jorik Nonnekes contributed to 1A, 1C, 2C, and 3B.

Michael X Cohen contributed to 1A, 2C, and 3B.

Helena Cockx contributed to 2C, and 3B.

Sabine Janssen contributed to 2C, and 3B.

Rick C. Helmich contributed to 2C, and 3B.

Richard van Wezel contributed to 1A, 1B, 2A, 2C, and 3B.

