## Appendices for "Relationship between eye movements and freezing of gait during turning in individuals with Parkinson’s disease"

### Appendix A. Workflow of the algorithm for quick phase detection

| Steps for analyzing EOG signals in our developed system | | Remarks |
|---|---|---|
| 1. Preprocessing | 1.1 Up-sample electrooculography signals from 500 Hz to 1000 Hz;<br>1.2 Low-pass filtering [< 30 Hz];<br>1.3 Subtract the baseline drift from the signals:<br>1.3.1 Decompose the signals using wavelet decomposition (WD) method with Daubechies 4 wavelet;<br>1.3.2 Get the baseline from the approximation outputs from WD method in level 10 (*figure S1* shows an example);<br>1.4 Median filtering with a 50-millisecond window; | To reduce interference caused by muscle or movement artefacts, etc.;<br>An example before and after preprocessing is shown in *figure S2*. |
| 2. Feature extraction | 2.1 Commonly used features in EOG analysis [1]:<br>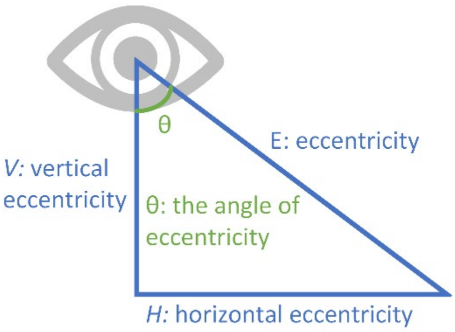<br><br>- Velocity of eccentricities:<br>$E_{velocity} = E_{position}(n) - E_{position}(n-1)$ with<br>$E_{position}(n) = E(n) - E(n-1)$, and<br>$n$ denotes each time sampling;<br>- Acceleration of eccentricities:<br>$E_{acceleration} = E_{velocity}(n) - E_{velocity}(n-1)$;<br>- Distance between the eccentricities at every second sampling points:<br>$E_{distance} = E(n) - E(n-2)$;<br>- Eccentricities in angular:<br>$\theta_{position} = \theta(n) - \theta(n-1)$;<br>- Angular velocity of eccentricities:<br>- $\theta_{velocity} = \theta_{position}(n) - \theta_{position}(n-1)$<br>2.2 Remove outliers of the features using interquartile rule;<br>2.3 Normalized features; | *1. König, S. D. & Buffalo, E. A. A nonparametric method for detecting fixations and saccades using cluster analysis: Removing the need for arbitrary thresholds. J. Neurosci. Methods 227, 121–131 (2014).* |
| 3. Clustering | 3.1 Use k-means clustering partitioned the data points into two clusters: quick phases and slow phases;<br>3.2 The cluster of quick phases has a higher average velocity, and the another cluster is of slow phases;<br>3.3 A series of continuous sampling points in the same cluster was treated as a phase (*figure S3* shows an example);<br>3.4 Refine the clusters:<br>3.4.1 The outliers in the cluster of slow phases are considered as quick phases;<br>3.4.2 Re-cluster (k-means) the current quick phases using main sequences: amplitude, duration, and peak velocity of the quick phases; | The refinement can improve the accuracy of quick phase detection. |

3.5 Check whether the refined quick phases are next to each other:

3.5.1 Merge the neighbor quick phases when they have same direction;

3.5.2 Remove the neighbor quick phase with a lower velocity when the two neighbor quick phases have different directions.

3.6 Get the time stamps of the final quick phases and treat the rest as slow phases (*figure S4* shows an example of the final detected quick phases).

Figure S1. An example of a 2-minute session signals and their baseline extracted by wavelet decomposition.

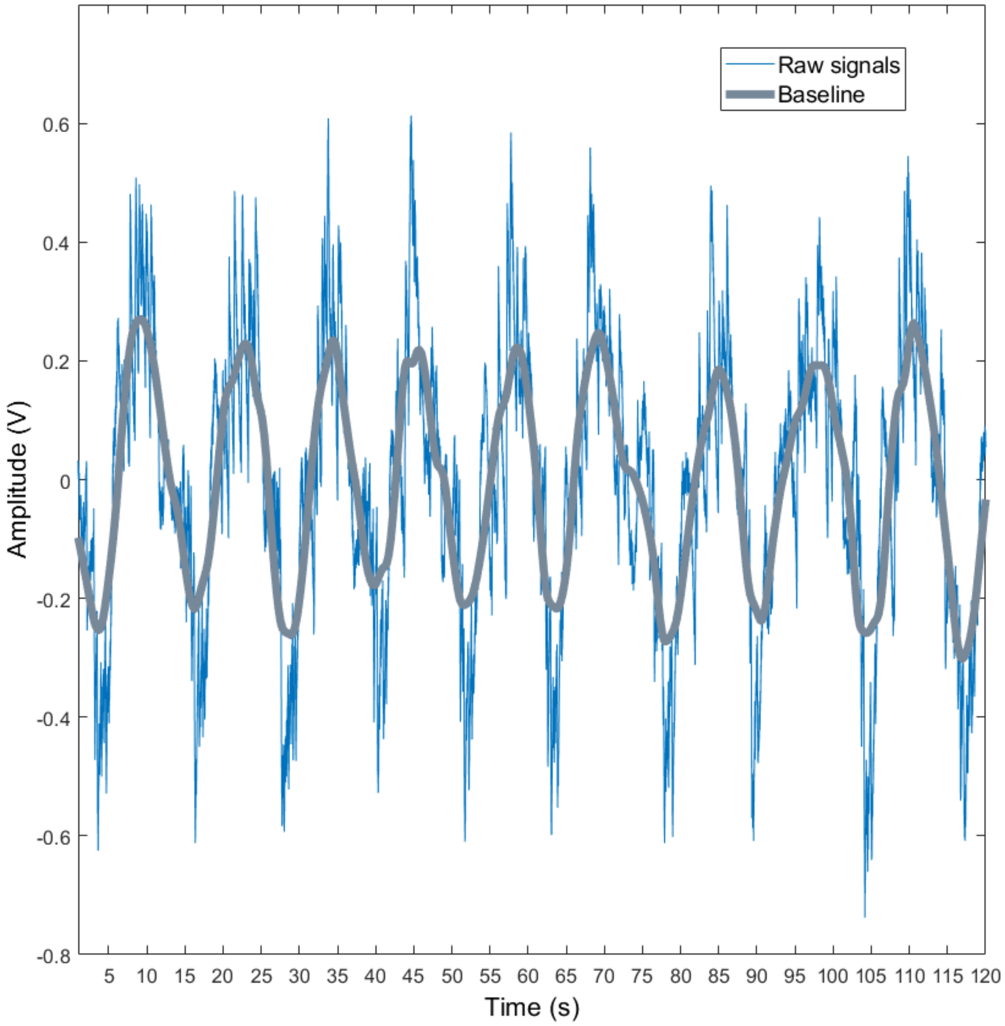

Figure S2. Horizontal EOG signals before and after preprocessing. (A) An example of a 2-minute session. (B) Close-up plot of the first 10 seconds in (A).

A

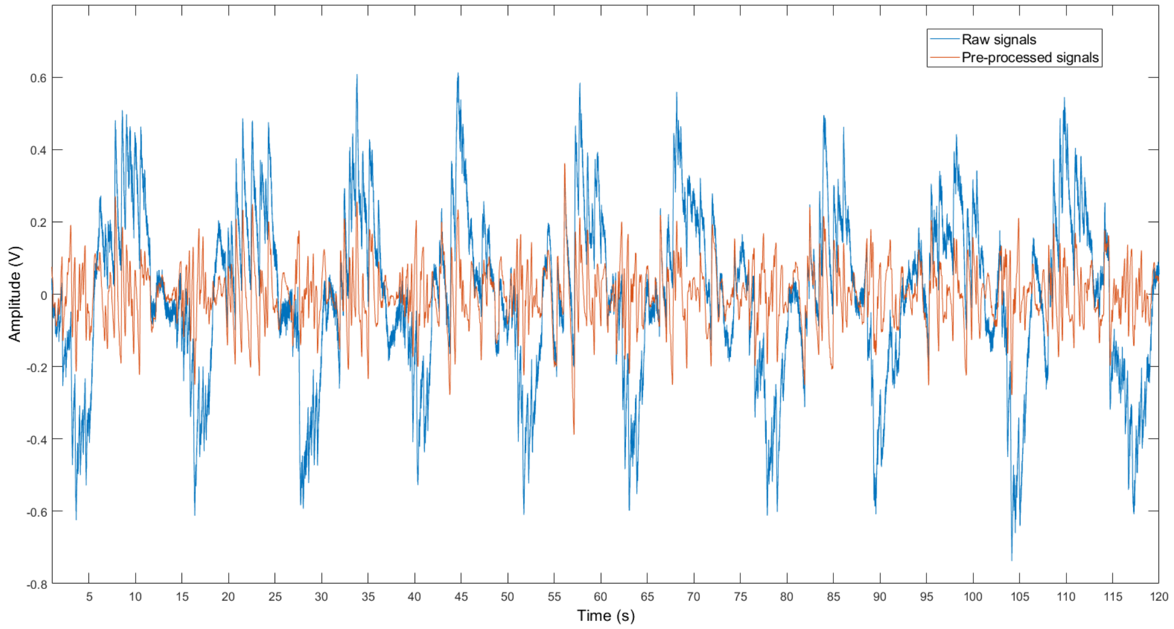


B

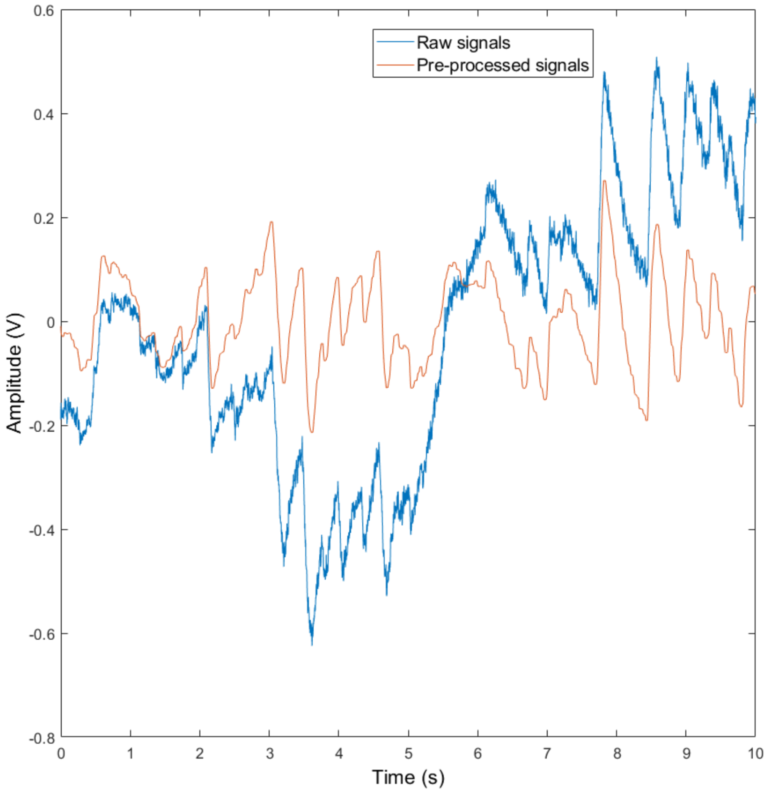

Figure S3. An example of quick phases detected in 10-second signals before the cluster refinement.

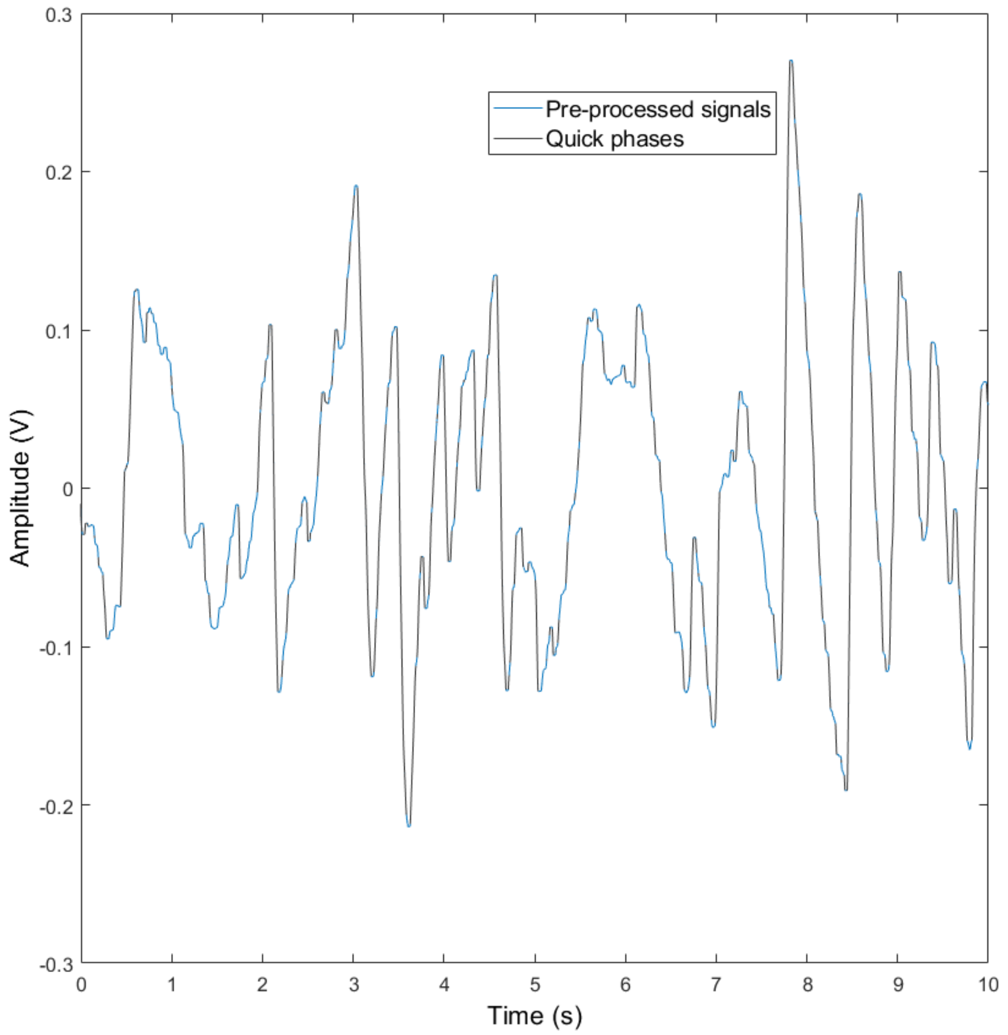

Figure S4. An example of quick phases detected in 10-second signals after the cluster refinement.

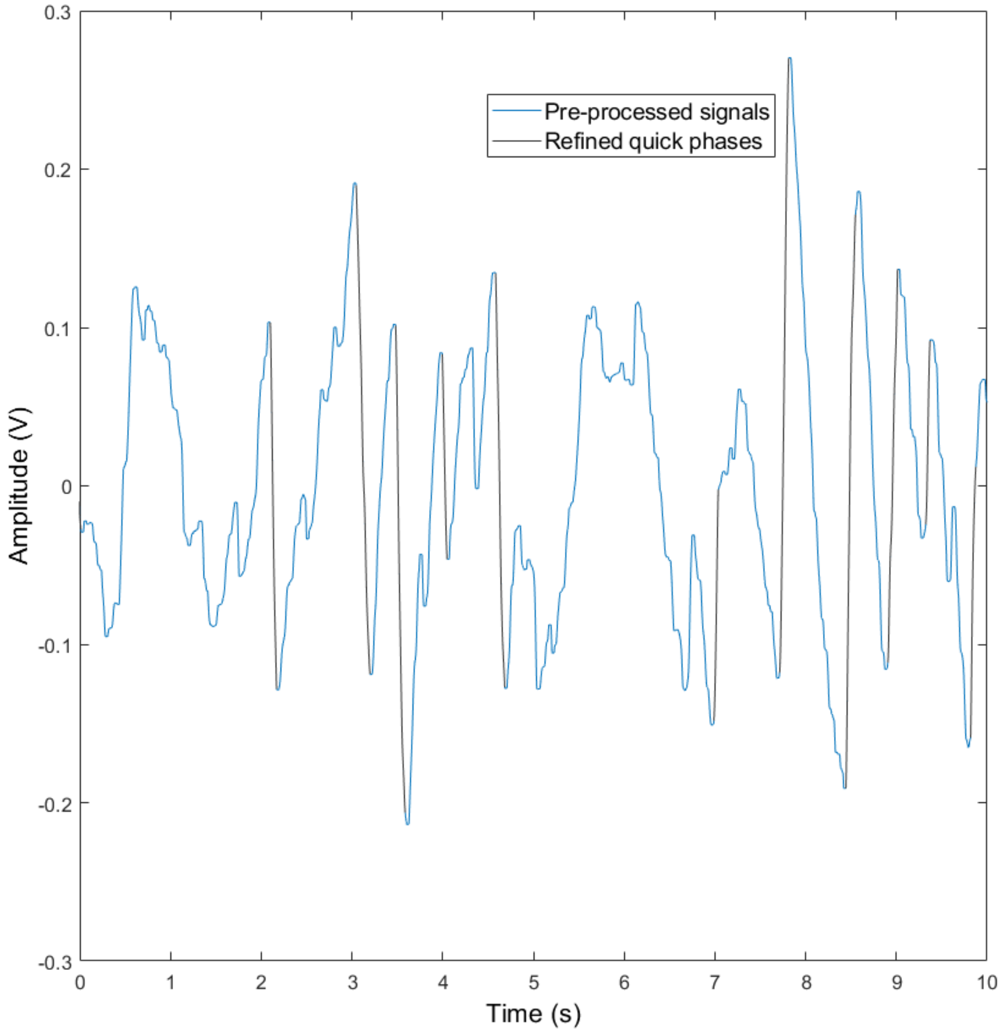
